# High Variation in Purity of Consumer-Level Illicit Fentanyl Samples in Los Angeles, 2023-2025

**DOI:** 10.1101/2025.06.27.25330446

**Authors:** Chelsea L. Shover, Adam J. Koncsol, Morgan E. Godvin, David Goodman-Meza, Bryce Pardo, Michelle Poimboeuf, Caitlin A. Molina, Ruby Romero, Jasmine Feng, Joseph R. Friedman

## Abstract

**Background:** The variation in purity of illicitly manufactured fentanyl has been theorized to be a key driver of overdose. However, data on the purity of illicitly manufactured fentanyl in the United States typically comes from law enforcement seizures and is rarely available at the consumer-level, which is most relevant to overdose risk.

**Methods:** Samples were analyzed from a community-based drug checking program operating at four geographic sites in Los Angeles County, California 2023 Q1 to 2025 Q2. Participants answered an anonymous survey about sample characteristics. Qualitative and quantitative analyses were conducted leveraging directly-observed mass spectrometry (DART-MS) and Liquid chromatography mass spectrometry (LC/MS) respectively. LC/MS quantified a panel of compounds including fentanyl and fluorofentanyl. Composite fentanyl purity was estimated by adding the percent mass of fentanyl and fluorofentanyl.

**Results:** A total of 353 samples had either fentanyl, fluorofentanyl, or both, quantified. Average purity was 10.0%, SD 11.1%, range 0.1%-64.9%. Samples expected to be fentanyl (n=308) had higher average purity (10.9%) compared to those expected to be heroin (n=24, average purity=2.7%) or other drugs. Powder samples (n=318) had higher average concentration (10.8%) compared to pills (n=11, 1.4%) or tar (n=22, 3.2%). Of expected-fentanyl samples, 42.5% (n=117) had a fentanyl purity of less than 5%, while 17.5% (n=51) had purity over 20%.

**Conclusions:** We found high variation in fentanyl purity among consumer-level samples sold as fentanyl, which may explain overdose among people with opioid tolerance. Fentanyl concentration was lower among samples sold as heroin, other drugs, or in pill form, and was particularly low among expected non-opioid drugs.

## Introduction

In the United States, the current “fourth wave” of the overdose crisis is characterized by deaths involving illicitly manufactured fentanyl (IMF) often mixed or used together with other psychoactive substances.^1^ Information about the purity of IMF has been limited, and mostly based on law enforcement drug seizure data.^2–4^ Quantitative testing of consumer-level samples of IMF is done regularly in at least one major Canadian drug checking program,^5^ but there are few reports in the scientific literature.^6,7^ The purity of consumer-level samples of IMF can be expected to differ from law enforcement samples, which are often wholesale quantities that have not yet gone through all the steps of adulteration that typically occur between producer and consumer.^8,9^ Understanding variation in purity and co-detected compounds is critical to responding to the evolving challenges that IMF pose to public health.

In 2023, the most recent year for which complete data are available, Los Angeles County led the United States in number of deaths involving “synthetic opioids other than methadone,” an *International Classification of Diseases 10*^*th*^ *Edition* code widely considered to be almost entirely comprised of IMF and other illicitly manufactured synthetic opioids.^1,10^ Relative to other areas of the U.S., Los Angeles was late to develop an IMF market, and deaths involving fentanyl have largely also involved other drugs such as psychostimulants.^11,12^ Attributing polysubstance deaths to unexpected fentanyl exposure (e.g., using cocaine or methamphetamine contaminated with fentanyl) versus intentional co-use (e.g., using a speedball or goofball) has been challenging. Prevention strategies for these two scenarios differ, and testing drug product samples stands to add some clarity.

In this analysis, we aimed to characterize samples of fentanyl submitted to a community-based drug checking program in Los Angeles, California. The primary objective was to describe the purity (concentration) of samples containing fentanyl, with secondary objectives to a) characterize the other components of these samples, and b) concentration variation across small geographic areas.

## Methods

### Sample and data collection

Using methods previously described, we leverage data from a community-based drug checking program, Drug Checking Los Angeles.^6^ Samples were submitted anonymously by community members for testing at sites throughout Los Angeles County between September 2023 and April 2025. Individuals providing drug samples for testing were invited to complete an anonymous, voluntary survey that asked what they expected the drug to be, the neighborhood where they obtained it, and other characteristics of the sample. Initial field testing was performed at the community sites with Fourier transform infrared (FTIR) spectrometer and immunoassay test strips, except when participants requested mail-in testing only. Laboratory testing was performed by the National Institute of Standards and Technology (NIST). All sample underwent qualitative analysis (identifying presence but not amount of over 1,300 different compounds), performed with direct analysis in real-time mass spectrometry (DART-MS).^13^ A subset of these samples were subject to quantitative testing using liquid chromatography mass spectrometry (LCMS).^6^ Study protocols were approved by the UCLA IRB (IRB-22-0760 and IRB-22-1273).

### Sample selection and preparation for LCMS

Samples sent for quantitative testing were scooped with a 5 mg microscoop into amber glass vials pre-filled with acetyl nitrile. Selection for quantitative testing was based on expected contents, field-test results, and availability of sample and vials (e.g. in instances where the field team had fewer vials than samples, samples provided after depletion of vials were not able to be quantified). All samples expected to be fentanyl, as well as those that tested positive on a fentanyl test strip or showed fentanyl on the FTIR, were sent for quantification provided there was sufficient sample to scoop and vials were available. Other samples that underwent quantification included those containing other compounds on the current quantitation panel, whether based on reported expected contents or field tests. Per protocol, samples that were expected to be a substance not included in the quantitation panel (e.g., benzodiazepines, phencyclidine, 3,4-Methylenedioxymethamphetamine (MDMA)) were only quantified if field tests showed evidence of unexpected compounds (e.g., fentanyl, methamphetamine).

### Quantitative testing

For every sample, the quantitation panel included: fentanyl, fluorofentanyl, two fentanyl precursors (4-ANPP, phenylethyl 4-ANPP), heroin, cocaine, methamphetamine, xylazine, medetomidine, and tetracaine. A subset of samples underwent testing with expanded panels in use by NIST during part of the study period (see supplemental table 1). Results were reported in percent mass, with 0.1% imputed as the limit of quantitation.

**Table 1.**
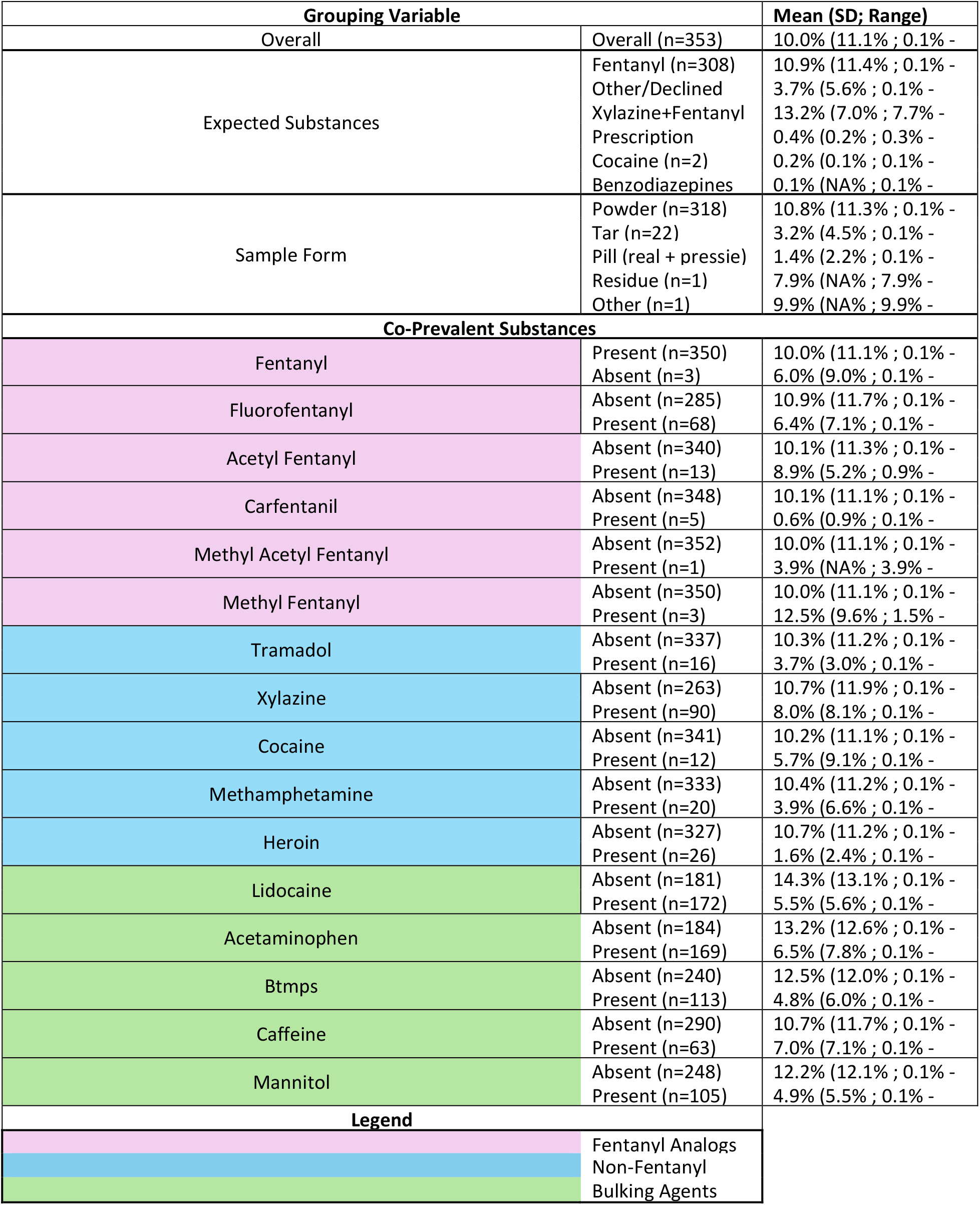
Fentanyl and Fluorofentanyl Concentration By Expected Substance, Sample Form, and Co-prevalent Substances. For all groups the concentration of fentanyl and fluorofentanyl is summed and treated as a continuous variable. This is shown overall, as well as broken down by expected substances, sample form, and a number of co-prevalent substances (present/absent according to DART-MS). Co-prevalent substances are sorted by class, which is shown by color.

This analysis includes all samples collected between 2023 Q3 and 2025 Q2 that underwent quantitative testing and were found to contain fentanyl or fluorofentanyl. This time period includes a period when people who use fentanyl reported declining quality and availability, and a documented decline in national and local overdose deaths. A subset of the included samples were collected in Downtown Los Angeles in close time and geographic proximity. These were collected in three sessions in Q3 2024, each two to four hours long, with samples obtained from the approximately one quarter square mile of area surrounding a temporary testing site.

Descriptive statistics were calculated overall and stratified into four a priori-selected categories (which generally align with 4 quartiles) to characterize very low (<1%), low (1-5%), medium (5-15%) and high (>15%) purity fentanyl. All analyses were performed using R Version 4.4.1.

## Results

In total, n=1,763 samples collected between 2023 Q1 and 2025 Q2 were analyzed. Of these, n=353 both underwent LC/MS testing and were found to contain either fentanyl (n=350) or fluorofentanyl (n=68), and were therefore included in this study. Only n=3 samples contained fluorofentanyl but did not contain fentanyl. Fentanyl percent by mass (purity) ranged from 0.1% to 64.9% with mean of 9.6% (standard deviation 11.0%). Fluorofentanyl purity ranged from 0.06% to 28.9% with a mean of 3.0% (standard deviation 5.9%). The mean percent mass of the sum of fentanyl and fluorofentanyl (referred to as ‘fentanyl purity’ moving forward) was 10.0% (standard deviation 11.1%), range 0.1% to 64.9%. (Table 1). Overall, a skewed—and highly variable—distribution of fentanyl purity was noted (Figure 1). A total of 17.0% of samples (n=60) had very low fentanyl purity (under 1%), 28.3% (n=100) had low purity (between 1-5%), 28.0% (n=99) had medium purity (between 5-15%), and 24.9% (n=88) had high purity (above 15%).

**Figure 1.**
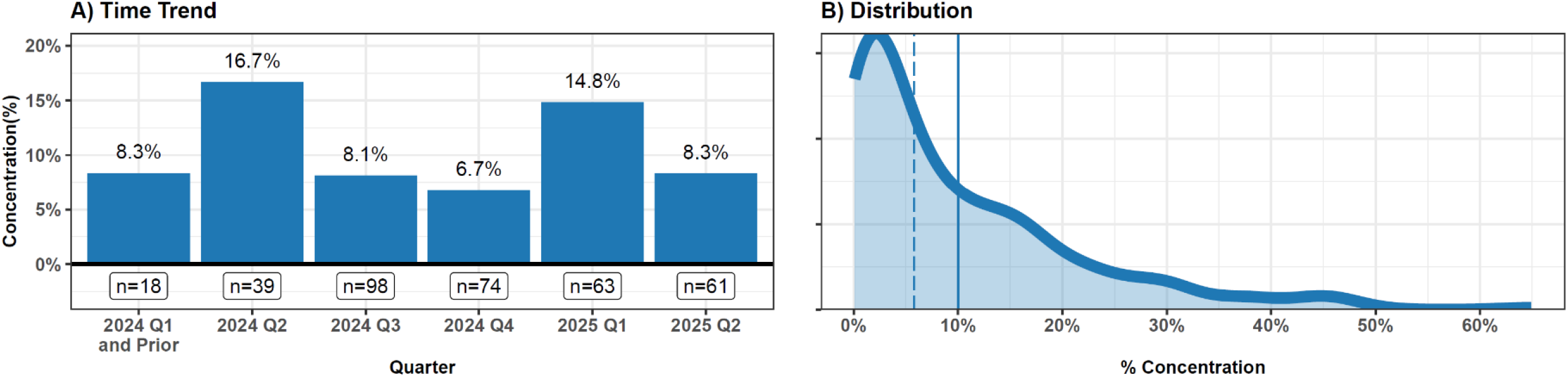
Fentanyl and Fluorofentanyl Concentration Among Fentanyl Positive Samples. For both plots the concentration of fentanyl and fluorofentanyl are summed and treated as a continuous variable. A) The time trend of average fentanyl and fluorofentanyl concentration is shown by quarter-year through 2025 Q2. Values prior to 2024 Q1 are grouped with 2024 Q1 due to small numbers. Sample size is shown under each bar. B) The overall distribution of fentanyl concentration (across all time points) is visualized using a kernel density estimator. The mean and median concentration are shown with a solid and dashed line, respectively.

The average purity varied from quarter to quarter, with a maximum average observed in 2024 Q2 (16.7%) and minimum observed in 2024 Q4 (6.7%) [Figure 1]. Nevertheless, linear regression results indicated that there was not a statistically significant, consistent positive or negative time trend (p=0.681).

The vast majority of samples (n=308) were expected to be fentanyl. These samples had higher average purity (10.9%) compared to those expected to be heroin (n=24, average purity=2.7%) or other drugs (Table 1). Fentanyl concentration was particularly low among samples expected to be prescription opioids (n=2, average purity 0.4%) cocaine (n=2,0.2%) and benzodiazepines (n=1, 0.1%). Of the n=353 quantified samples that contained fentanyl/fluorofentanyl, n=318 were powder, n=22 were tar, and n=11 were pills (n=10 expected to be opioids, n=1 expected to be a benzodiazepine). Powder samples had higher average fentanyl purity (10.8%) compared to pills (1.4%) or tar (3.2%).

Although fentanyl purity was generally higher among samples that did not contain other co-detected substances, among the subset containing both fentanyl and other drugs, there was generally a positive relationship between fentanyl purity and the purity of other quantified substances (Figure 2). All 6 substances that were quantified among a large enough sample for assessment (heroin, BTMPS, xylazine, methamphetamine, lidocaine, 4ANPP) showed highly skewed distributions, with positive correlations to fentanyl purity.

**Figure 2.**
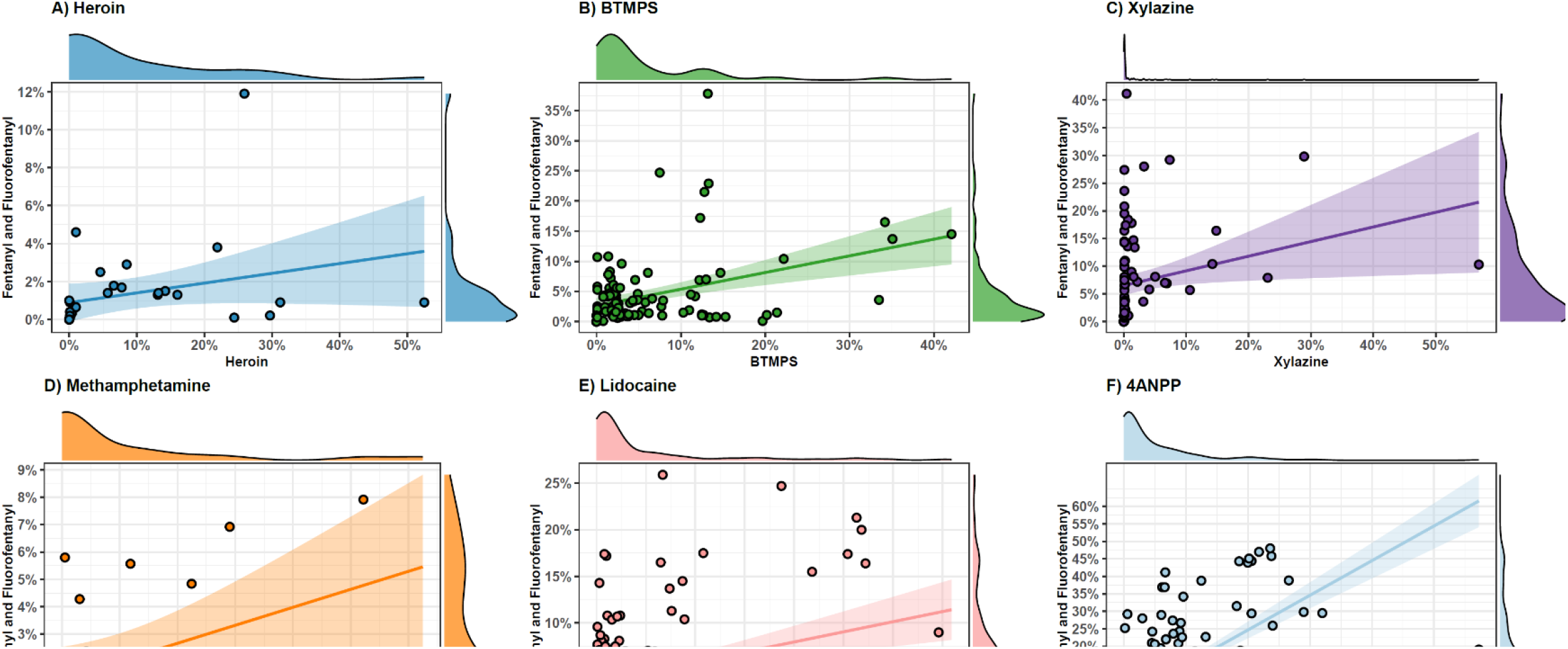
Concentration of Fentanyl and Fluorofentanyl Compared to Concentration of 6 other Commonly Co-Detected Substances. For all plots the concentration of fentanyl and fluorofentanyl are summed, treated as a continuous variable, and plotted against the concentration of 6 other commonly co-detected substances. For each, a linear relationship is graphed using OLS regression, and a 95% confidence interval of the relationship is shown. For each bivariate relationship, the distribution of both variables (fentanyl+fluorofentanyl, and the other substance shown) is visualized using a kernal density plot on each margin.

Fentanyl concentration varied by presence of other co-detected substances (Table 1). Samples that were fluorofentanyl-positive tended to have a lower average combined purity (6.4%) compared to fluorofentanyl negative samples (10.9%). A similar trend was seen for xylazine positive samples (8.0% vs 10.7%), BTMPS (4.8% vs 12.5%), cocaine (5.7% vs. 10.4%), methamphetamine (10.4% vs 3.9%) and heroin (10.7% vs. 1.6%). Across these substances, the presence of a contaminant was associated with lower fentanyl concentration. Similarly, co-detected substances differed by purity category (Figure 3). Lidocaine, BTMPS, heroin, xylazine, methamphetamine, and cocaine were less common among high purity fentanyl samples.

**Figure 3.**
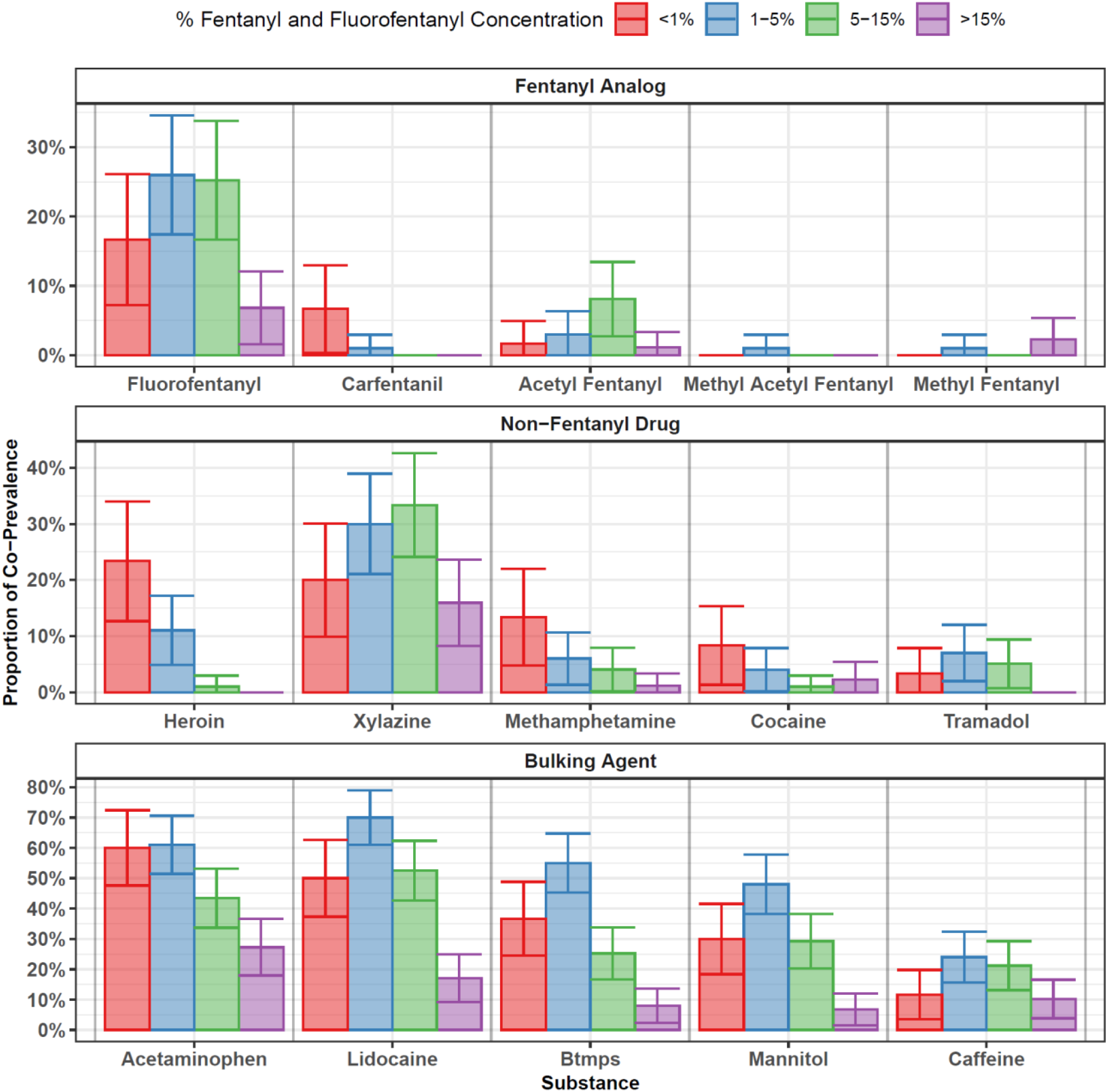
Co-Prevalence of Selected Substances by Fentanyl and Fluorofentanyl Concentration Category. The concentration of fentanyl and fluorofentanyl is summed and treated as a categorical variable with four levels. The co-prevalence (from 0-100%) is shown for a number of selected substances (according to present/absent status from DART-MS) separate for each categories of fentanyl concentration. The four categories roughly correspond to quartiles of fentanyl + fluorofentanyl concentration. 95% confidence intervals around each percentage are shown with vertical bars

In an exploratory case study analysis of samples collected in a single day, from a small geographic area, we observed very high variability in fentanyl concentration and other additives. (**Figure 4**). For instance, of the n=13 total samples, n=5 had a combined fentanyl concentration of about 50%, n=4 had a fentanyl concentration of about 10-20%, and n=4 had a concentration of less than 5%. N=3 samples had a considerable proportion of BTMPS. N=5 had lidocaine. N=1 sample had the majority of its combined fentanyl concentration represented by fluorofentanyl. This illustrates the chaotic and unpredictable nature of the illicit drug supply in Los Angeles, even in the same microneighborhood, on the same day.

**Figure 4.**
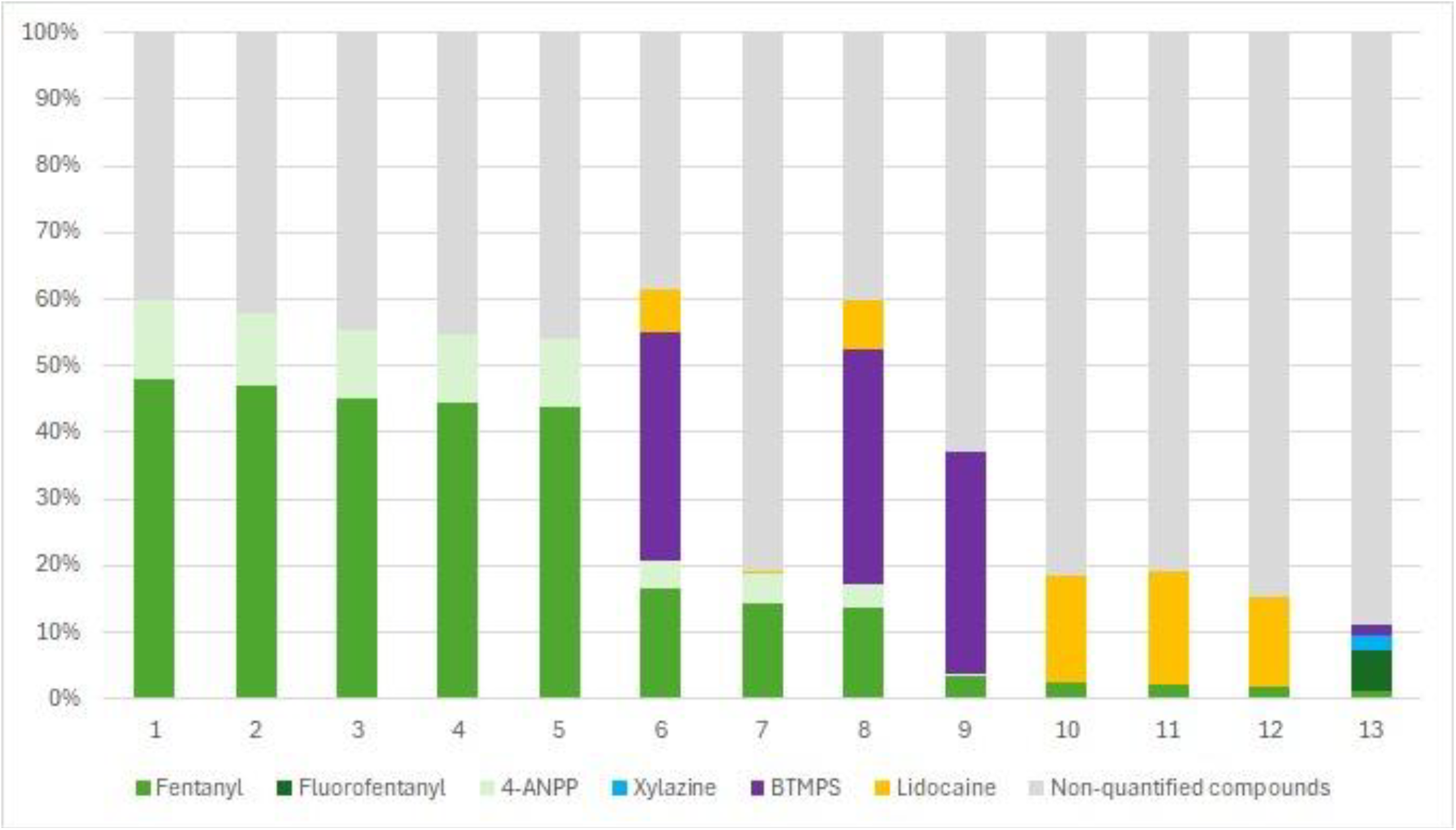
Quantitative Testing Results of Fentanyl Samples Collected in Close Geographic and Temporal Proximity. The 13 samples shown above were collected on a single day over the course of three hours from a geographic region in Downtown Los Angeles of approximately 10 square blocks. All samples were powder sold as fentanyl.

## Discussion

In a study of drug product samples submitted for quantitative testing at a community-based testing program in Los Angeles County, we found a large range of variability in fentanyl purity. Over half of the tested samples contained less than 5% fentanyl by mass, while a substantial minority had over 15%, reaching as high as 65%. In subsets of samples intentionally collected in small geographic and temporal proximity, the strongest purity of a white powder sold as fentanyl exceeded the weakest by 24-fold. This variation highlights how overdose risk changes from batch to batch. To our knowledge, detailed quantitative testing of consumer-level samples of fentanyl has previously been limited in the scientific literature. As such, these data represent a rarely seen look into what specifically is in drugs purchased as “fentanyl”.

Samples presumed to be fentanyl (rather than heroin or other opioids) generally had larger amounts of fentanyl detected, and we found that co-detected compounds varied by fentanyl purity category. Generally, higher purity samples were more likely to have various fentanyl synthesis chemicals detected, while lower purity samples were more likely to contain compounds typically viewed as adulterants that may plausibly have clinical health effects, such as local anesthetics, acetaminophen, and the UV stabilizer BTMPS. The range of concentration highlights how overdose risk may differ tremendously between batches obtained in very close geographic proximity. While lower purity fentanyl may carry reduced risk of fatal overdose compared to higher purity fentanyl, the presence of adulterants—which could have additional clinical effects—presents avenues for further inquiry.

The results underscore the need for expansion of quantitative drug testing in settings with IMF to understand what consumers are using. This strategy may be particularly impactful as a tool to support preparation strategies in settings with emerging or pre-emerging IMF markets, e.g. Western Europe.

Testing for fentanyl using immunoassay test strips has been widely practiced as a harm reduction strategy, but these tests indicate only the presence and not the amount of fentanyl. As a population-level public health strategy, fentanyl test strips can sound the alarm that fentanyl has arrived in a given drug market, but quantitative testing can better characterize the extent to which it is an adulterant versus a main product or “drug of choice.” Quantitative community-based drug checking may also represent an important community-led surveillance tool to improve public health. Moreover, such initiatives offer opportunities to engage with people who use drugs about various aspects of the illicit market to better characterize price, emerging substances, and other qualitative phenomena.

### Limitations

While the inclusion of quantitative testing is novel, the fact that samples came from only one metropolitan area and are inherently a convenience sample (i.e., what people choose to and are able to bring for drug checking) may limit the generalizability even within that metropolitan area. The samples were mostly powder rather than pills, and as such the findings likely do not capture the scope of pressed pills available on the illicit market.

In terms of testing methods, for most of the study period only two fentanyl analogues were quantified (fentanyl and fluorofentanyl). While these did represent the most common analogues, it is possible that not quantifying other analogues lead to an underestimate of effective fentanyl purity in these samples. However, the impact of this on the present analysis is likely small, given that only 2% of all samples had a fentanyl analog other than fentanyl or fluorofentanyl. It is also the case that different analogues have different strengths, so adding together fentanyl and fluorofentanyl may not reflect the total potency of the fentanyls in the drug, particularly since different fluorofentanyl isomers have different potency.^14^ Given the dominance of traditional fentanyl in the Los Angeles market, the impacts of this source of biad on the analysis are likely relatively small.

In conclusion, we found extreme variability in the purity of fentanyl in consumer-level samples, often in samples obtained within the same small geographic area on the same day. This variation may partially explain overdose risk among people with opioid tolerance, and may point the way for community-led drug supply surveillance efforts to understand risk and harm for people who use opioids. Findings about differing co-detected compounds may suggest avenues to study different synthesis methods and manufacturing decisions (i.e. using different diluents to bulk up a sample). Quantitative testing of illicitly manufactured drugs is poised to make unique contributions in understanding and responding to changes in the illicit drug supply that ultimately impact public health.

## Data Availability

Data used in this analysis are sensetive and cannot be shared in raw form. However aggregate statistics can be provided by the authors upon reasonable request.

## Acknowledgments

We gratefully acknowledge our partners at the National Institute of Standards and Technology who conducted laboratory testing as part of the Rapid Drug Analysis and Research (RaDAR) program.

